# Partial lockdown on unvaccinated individuals promises breaking of fourth COVID-19 wave in Bavaria

**DOI:** 10.1101/2021.11.28.21266959

**Authors:** Tobias Krebs, Michael J. Moeckel

## Abstract

**Purpose of this report:** The aim of this rapid communication is a projection of the development of the fourth COVID-19 wave in the federal state of Bavaria in Germany, taking into account different lockdown scenarios especially for unvaccinated individuals. In particular, the number of infections and the occupancy of intensive care facilities are considered.

**Applied Methods:** We use the agent-based epidemiological simulator Covasim for discussing various epidemiological scenarios. Firstly, we adapt and calibrate our model to reproduce the historical course of the COVID-19 pandemic in Bavaria. For this, we model and integrate numerous public health interventions imposed on the local population. As for some of the political actions rigorous quantification is currently not available, we fit those unknown (free) model parameters to published data on the measured epidemiological dynamics. Finally, we define and analyse scenarios of different lockdown scenarios with restrictions for unvaccinated individuals in different areas of life.

**Key message:** The results of our simulations show that in all scenarios considered, the number of infections, but also the number of severe cases, exceed previous maximum values. Interventions, especially restrictions on contacts of unvaccinated persons, can still mitigate the impact of the fourth COVID-19 wave on populations substantially. Excluding unvaccinated students from attending classes has only a small impact on the public health burden. However, many severe cases can be prevented by reducing community and/or work related contacts of unvaccinated people, e.g, by achieving high home office rates.

## Background

The COVID-19 pandemic has kept the world on suspense for almost two years and and the number of deaths associated with COVID-19 recently rose to over 100,000 (Robert Koch Institut 2021a). After a summer with low infection rates, the Robert Koch Institute has recorded a rapidly increasing number of infections since the beginning of the cold season, which now exceed all previously measured statistics (Robert Koch Institut 2021a, 2021c). The rate of fully vaccinated individuals in the population of Germany at the time of writing this report was 68%, in Bavaria slightly lower at 66% (Robert Koch Institut 2021b). A considerable fraction of the German population is -despite the availability of sufficient vaccine-unwilling to be vaccinated for a variety of reasons (Graeber et al. 2021) and thus still represent a large reservoir of susceptible individuals for corona virus SARS-CoV-2 infection. At the time of this report, rising numbers of infections have raised concern on the resilience of the healthcare system to cope with the number of expected hospitalizations. Therefore, the public debate has returned to defining suitable public health interventions to reduce the number of severe cases. Since unvaccinated individuals are at much higher risk of developing severe conditions, protecting them from infection becomes a cornerstone of a public health strategy. This report contributes to the debate by investigating the effectiveness of a partial lockdown, i.e. contact restrictions, only within the sub-population of unvaccinated individuals. Our results show that among the considered scenarios such partial contact restrictions emerge as the most effective way to protect unvaccinated populations from infection and to reduce the peak load on the healthcare system..

Since the beginning of the pandemic, epidemiologists have been using computer simulations in an attempt to meet the demands of decision-makers for scientific assessment of political options and forecasts on the development of the pandemic. A variety of models with different approaches have been adapted or newly developed for the COVID-19 pandemic (Panovska-Griffiths et al. 2021). Agent-based models have proven to be capable of representing the complexity of the pandemic in some detail, including public health interventions, such that the number of available models grew rapidly (Lorig et al. 2021). Covasim (Kerr et al. 2020) is one of these agent-based COVID-19 simulators that have already been used by various scientists around the world for epidemiological simulations (Stuart et al. 2020; Latkowski and Dunin-Kęplicz 2021; Scott et al. 2021); we adapted it for the federal state of Bavaria in Germany.

## Methods

All our results are based on the agent-based simulation tool Covasim, which we used to simulate and compare different lockdown scenarios for the federal state of Bavaria in Germany. We created a synthetic population that matches statistically the real population of Germany in essential aspects, such as age structure or household composition. Since simulations for the 13.1 million inhabitants of Bavaria would take an extremely long time, we decided to scale up from a reduced sample. Thus, we carried out our simulations with 71,000 agents and all absolute numbers were scaled accordingly by a factor of 185. Contact networks between the agents have been set up for four typical environments: home, school, work and free time. The simulation essentially calculates probabilities of virus transmission from one agent to another given existing contacts, allowing for a variety of active countermeasures. In Covasim, a stochastic procedure determines actual infection events for each day by combining specific transmission probabilities and contact networks. Thus, Covasim provides a comprehensive picture of the infection incidence in a population and depicts the temporal evolution of the epidemiological dynamics.

We integrated non-pharmaceutical (public health) and pharmaceutical (vaccinations) interventions applied in Bavaria into the Covasim simulator and, whenever possible, quantitatively modelled their extent using publicly available data. Whenever the regulations in Bavaria were not uniform, we applied the regulations of the city of Aschaffenburg for the federal state. The following aspects were accounted for into our model:

- We calibrated the base transmission probability such that actions like the obligation to wear facemasks, increased hygienic barriers or enlarged average spacing of people at public places are included. From late summer 2021 onwards, due to declining compliance in the population with some regulations, a readjustment of the baseline transmission probability was necessary, reflecting the resulting increased probability of infection in the case of contact between two agents, in order to reproduce the real pandemic trajectory.
- The crossover from the wild variant of COVID-19 to alpha and delta variants were modelled continuously as the simulation progresses
- PCR testing of the population is performed daily. We did not apply a uniform probability for selecting agents for testing; however, we ensured that infected individuals with symptomatic courses are more likely to receive testing than asymptomatic cases or uninfected individuals. Especially all severe and critical cases are tested immediately. Covasim sends all diagnosed agents to quarantine. Estimates have been made for the number of tests to be conducted in the future, based essentially on the assumption that approximately the same number of tests will be conducted as last year.
- At the same time, we modelled contact tracing by public health departments, which trace direct contacts of infected individuals and send them to quarantine.
- We included full school closures as well as hybrid teaching models with only half of the students present at school.
- We simulated “working from home”-arrangements by assuming a reduced number of contacts at the work level and represented private contact restrictions by a reduced number of contacts at leisure activities.
- During the summer months (until end of September) there is a reduced risk of infection (so-called “summer effect”), possibly because of a shift of contacts from indoor to outdoor locations. We modelled it by a correction to the risk of infection.
- We simulated travel during summer vacations by randomly imported infections during the vacation season.
- Vaccinations, including first, second and booster vaccinations, were distributed to the population according to the official figures of the Robert Koch Institute. We made assumptions for the number of future vaccinations. For simplicity, we assumed that the Biontech/Pfizer vaccine was used for all vaccinations.
- Starting on December 01, 2021, additional partial lockdown measures were simulated, which particularly affect different areas of life of unvaccinated persons.

To implement the described measures in our Covasim model, some adjustments to the software were necessary. In particular, changes were made to the implementation of vaccinations, for example to enable booster vaccinations (Krebs et al. 2021) or to distribute them to certain age groups according to daily changing quotas. Further source code modifications were necessary to integrate available data.

Procedures are described in the literature to allow for an automatic calibration process of agent-based models (Hazelbag et al. 2020). In this case, however, manual adjustment of the model parameters proved to be the best way of fitting. When calibrating the model, we fixed the free parameters of the model in such a way that the simulated curves and the provided real data of the 7-day incidence and the critical cases in the period from February 01, 2020, to November 24, 2021, visibly matched well. Then, we used the calibrated model of the pandemic as a starting point for simulating the following future lockdown scenarios:

- **Scenario NI, “no interventions”**: This scenario represents the case where no further action is taken. All contacts remain as they were in October and November of 2021.
- **Scenario L2020, “lockdown 2020”**: Very strict interventions were imposed in March 2020 to mitigate the impact of the first COVID wave. In this scenario, analogous to the 2020 lockdown, all schools were closed; work contacts were reduced by 50% and leisure contacts by 70%. This scenario provides a reference of how much could be achieved with a hard lockdown.
- **Scenario PL S100, “partial lockdown at school”**: In this scenario, it is simulated that only fully vaccinated students are allowed to attend classes on site in the schools and unvaccinated students are taught via home schooling.
- **Scenario PL W100, “partial lockdown at work”**: Another possibility for a lockdown for unvaccinated people would be to send this group of people (partially or completely) to the home office. Here we consider the case where all unvaccinated workers stay at home.
- **Scenario PL W50&C50, “partial lockdown at work and leisure time”**: Interventions that affect schoolchildren are very unpopular. Moreover, it will hardly be possible to let all unvaccinated workers work from home. Therefore, in this last scenario, we consider the more realistic case where 50% of the unvaccinated workers work from home and at the same time, all unvaccinated persons reduce their community contacts by 50%. We generate each simulation output from an ensemble of 200 numerically equivalent implementations for the same parameter values to account for Covasim’s stochastic approach. Then, we present results as ensemble averages to reduce fluctuations and include statistical uncertainties.

## Limitations

The results of this report are to be considered preliminary and may change with further data and findings collected in the future. In particular, only findings and local data of Bavaria or Germany have been included. Little data is available on the actual implementation of public health orders. Therefore, we had to refer to plausible assumptions or fitted parameters for modelling some public health interventions. The stability of our results with respect to changes in those assumptions has not been established in all cases. In principle, modified assumptions may have some influence on the simulations performed.

The greatest uncertainty of our simulation lies in the actual human behaviour. The degree of compliance is difficult to measure, but has a non-negligible impact on the evolution of the pandemic. Past data was used to calibrate, among other things, a baseline probability of transmission of the virus in a contact between two agents. This probability includes aspects such as wearing masks, following distance rules,etc. We did not try to predict any behavioural changes from November 2021 until February 2022 but kept all related simulation parameters constant.

Furthermore, we also assume constant simulation parameters referring to other aspects in this study. For example, the emergence of new virus variants with modified infection probabilities could significantly influence the evolution of the pandemic.

## Results

Fig. 1 shows the result of calibrating our Covasim model to the course of the pandemic in the period from the beginning of the pandemic in March 2020 to the end of November 2021. Essentially, the simulation was able to reproduce the real course sufficiently well. Both the first three Covid 19 waves and the beginning of the fourth wave were captured by our model. In this way, the previous history of the population, especially regarding existing immunisations through infections and vaccinations, is taken into account for the future scenarios.

**Fig. 1:**
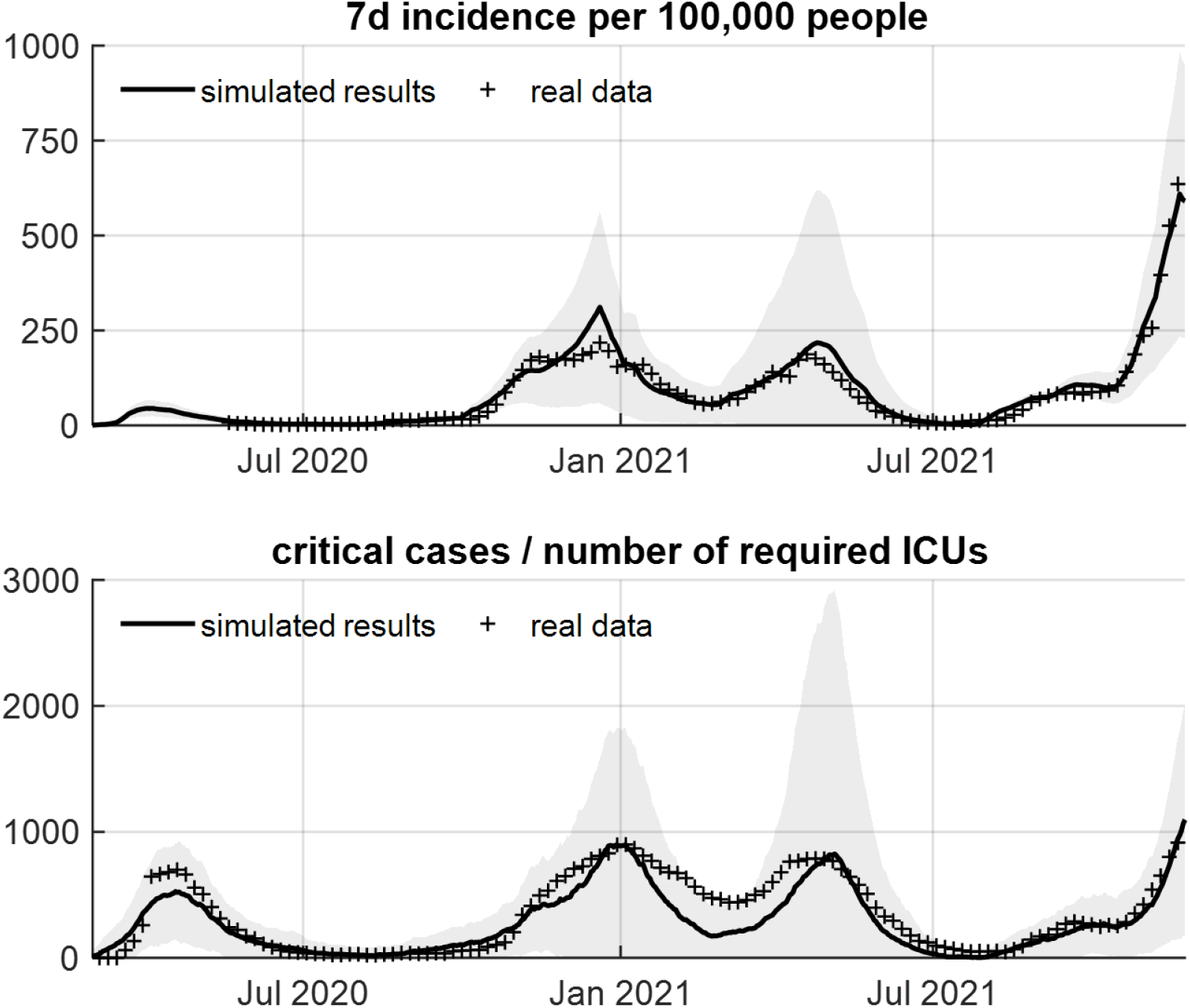
The two plots illustrate the calibration of the Covasim model. The solid line represents the results of the simulations and the marked dots represent real aggregated data for Bavaria. of Simulation results and real data show reasonably good agreement for 7-day incidence and the number of critical cases requiring intensive care.

Fig. 2 shows the 7-day incidence and the number of critical cases requiring intensive care for the five scenarios considered. The time period of the figure covers one year from the beginning of March 2021 to the end of February 2022, so that a comparison with the third COVID 19 wave, which was also the first wave with the delta variant, is possible.

**Fig. 2:**
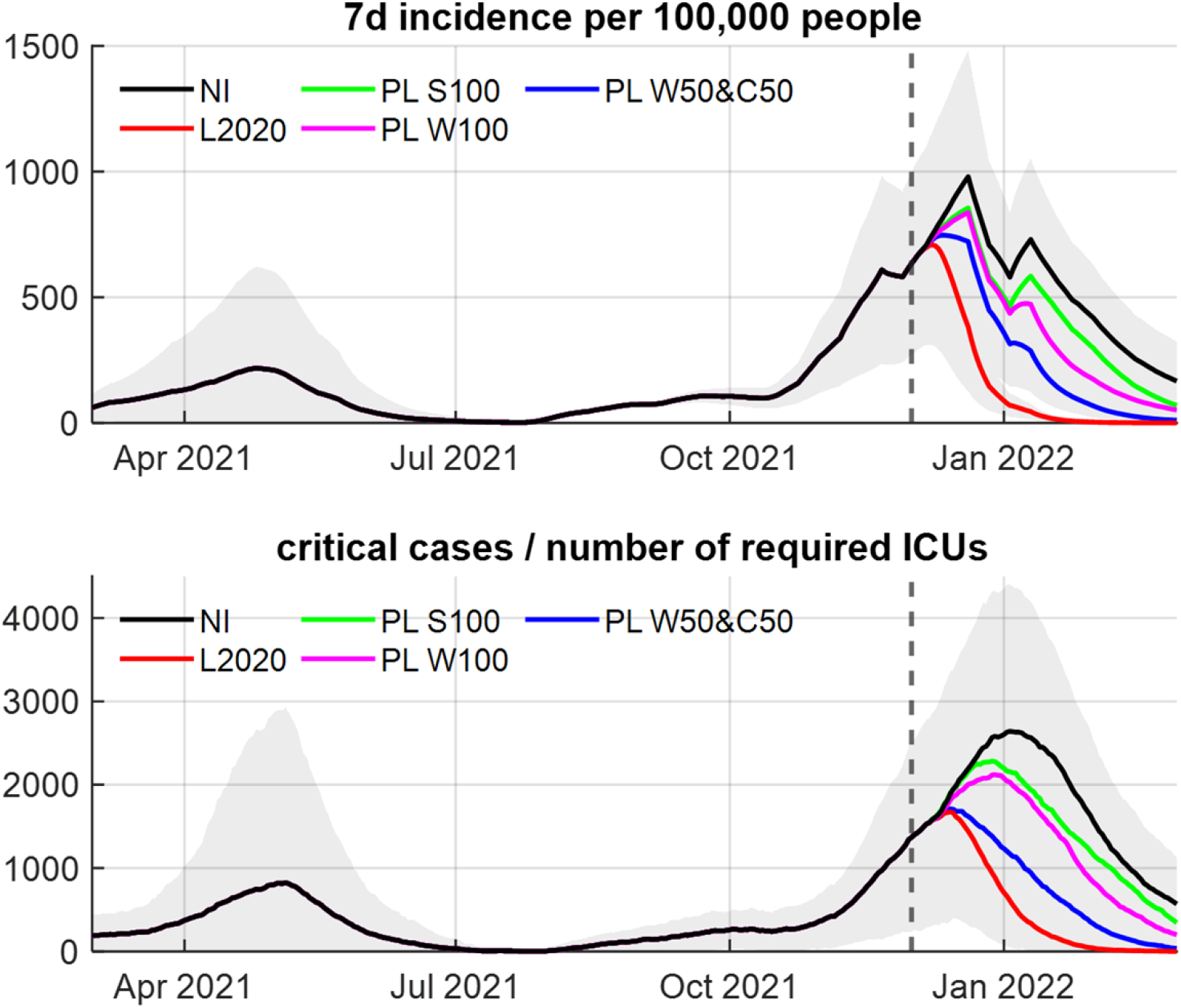
The simulation results for the 7-day incidence and number of intensive care units required are shown. The individual scenarios are colour-coded. Furthermore, the starting point of the interventions, 1 December 2021, is also marked with a dashed line.

## Discussion and Conclusions

At the beginning of all scenarios, the health system was already under strain due to many infections and critical cases.

The Scenario NI represents the worst-case scenario if no interventions limit the contacts of the population. In this case, the model projects a 7-day incidence of just under 1000 in the second last week of 2021 for the federal state of Bavaria and a total of more than 2600 intensive care units will be required in the first week of January 2021. This is almost three times the previous peak value during the first three waves.

However, the simulations also show that interventions starting on 1 December 2021 are able to mitigate the course of the fourth wave substantially. As the number of critical cases follows the increasing number of infections with some delay, a further short-term increase of critical cases is still to be expected even after interventions will come into effect. Scenario L2020 represents an extensive lockdown for the entire population by re-activating previously (March 2020) applied interventions and is regarded as a reference. Applying these interventions to the current situation, the 7-day incidence will reach about 700 and the number of critical concurrent cases will peak at roughly 1700. The values of the three other scenarios considered lie between Scenario NI and Scenario L2020.

The exclusion of unvaccinated students from classes in schools proved to be the least effective of the measures considered. Working from home of all unvaccinated workers was found to be somewhat more effective. According to our Scenario PL W50&C50, a very good mitigation of the fourth wave can be achieved by a combination of working at home and restricting leisure contacts of the unvaccinated population. Here, the maximum number of intensive care beds required is only slightly higher than in the hard lockdown of Scenario L2020. The total number of infections could also be significantly reduced in our simulation by the two measures described. Scenario PL W50&C50 thus also shows that no additional restrictions on the vaccinated population are required for a drastic mitigation of the fourth wave.

However, it is also noteworthy that even without further interventions in Scenario NI, the number of infections decreases shortly before the turn of the year and, somewhat later, the number of critical cases also decreases. We attribute this fact to the ever-increasing immunisation of the population, both through further vaccinations and through infection. As a result, the population’s immunisation is progressing towards herd immunity, making infections less probable.

However, our simulations show that decisive interventions can drastically reduce the impact on the health system, both in terms of peak burden and in terms of the number of cases that need to be treated, while not further restricting the vaccinated population.

## Data Availability

Only data from publicly available sources has been used.

## Acknowledgements

The authors gratefully acknowledge funding by the EpiL**AB**^KI^ project through the Bavarian Programme for Applied Research and Development at Universities of Applied Sciences (6^th^ funding period).

